# Promoting physical activity in glioma patients: insights from Dutch healthcare professionals

**DOI:** 10.1101/2025.09.29.25336660

**Authors:** Marieke E. C. Blom, Maxine Gorter, Philip C. de Witt Hamer, Johanna M. Niers, Judith G.M. Jelsma, Martin Klein, Linda Douw

## Abstract

**Background:** Although physical activity may play a significant role in enhancing quality of life for glioma patients, its integration into clinical care remains underexplored. This study assessed the experiences, perspectives, and barriers of healthcare professionals in promoting physical activity for glioma patients.

**Methods:** An online survey with 19 questions was distributed to Dutch healthcare professionals registered with the Dutch Neuro-Oncology Society (LWNO). Additionally, professionals were individually asked to remind and invite their colleagues to participate. Participants provided informed consent.

**Results:** Fifty-five professionals from 20 centers completed the survey, mainly neurologists (35%) and nurses (33%). Most professionals (58%) indicated receiving frequent questions about physical activity, particularly regarding safety and appropriate activities. Additionally, 76% stated they often proactively provide advice, typically recommending low-intensity activities like walking. Key barriers to making such recommendations included limited information materials (37%), knowledge (33%), and referral options (27%). Professionals’ concerns about physical activity included risks of overexertion and patient discouragement if activity would prove too challenging. Despite these challenges, 89% supported integrating physical activity into glioma care, and 56% expressed a need for specific guidelines. While professionals believe in the benefits for symptom management and quality of life, 62% were neutral about the strength of supporting evidence.

**Conclusions:** Physical activity is frequently discussed in glioma care, but recommendations are mostly inappropriate due to limited knowledge, resources, and guidance. Existing guidelines are rarely applied in practice. Future research should accumulate evidence, develop tailored guidelines, and equip professionals with tools to truly integrate physical activity into practice.

**Key points:**

- Physical activity is discussed in glioma care, but recommendations are challenging
- Barriers included limited informational materials, knowledge and referral options
- There is strong support for integrating physical activity

**Importance of the study:** Quality of life is a major concern for glioma patients, and physical activity has emerged as a promising complementary intervention to alleviate symptoms and enhance quality of life through its positive effects on physical and mental well-being. Although healthcare professionals recognize its value and frequently discuss physical activity with patients, significant barriers persist, including limited informational materials, insufficient knowledge, and uncertainty about referral options. While the benefits of physical activity are well-established in other types of cancer with cancer-specific guidelines, such guidelines and physical activity recommendations are not implemented in glioma care. Given the lack of glioma-specific evidence on the effects of physical activity, it remains unclear whether additional guidelines should be developed at all. This study identifies both the practical challenges and the evidence gap, highlighting the urgent need for targeted research, better resources, and structured support to help professionals effectively guide glioma patients in their physical activities.

## Introduction

Quality of life is a key concern for many cancer patients, including those with gliomas. Treatment of glioma often involves surgery, followed by radiotherapy, chemotherapy, or a combination of both. While patients may experience a period of stable disease after treatment, recurrence is virtually inevitable, making long-term well-being an ongoing challenge. Throughout the disease trajectory, patients often experience symptoms like fatigue, cognitive deficits, emotional distress, and reduced physical fitness that negatively affect their quality of life ^1-4^.

Physical activity has emerged as a promising complementary intervention to help improve symptoms and therefore quality of life in cancer patients, as it offers numerous benefits for both physical and mental well-being. Studies in various cancer populations have demonstrated improvements in physical fitness, muscle mass density, sleep quality, fatigue, dyspnea, and overall quality of life ^5-7^. However, most of this evidence is derived from patients with cancer not involving the central nervous system. Although research in glioma patients specifically remains scarce, it generally indicates that physical activity in these patients is both feasible and safe ^8-13^, even in the context of unique challenges such as neurological impairments, epilepsy, and disease-related fatigue ^3^. Besides being feasible and safe, physical activity also shows promising effects such as improvements in patient-reported outcomes^8-13^.

Despite these promising findings, the majority of glioma patients maintain low levels of physical activity throughout the course of their disease ^14-17^. The reasons for this inactivity are unknown. Furthermore, glioma patients often exhibit reduced physical fitness and physical functioning ^18-21^, which may be exacerbated by insufficient physical activity. This creates a negative cycle in which reduced physical fitness leads to even lower activity levels, underscoring the importance of regular physical activity for maintaining and improving physical fitness ^22^.

International guidelines, such as those from the World Health Organization (WHO) ^23^ and the American College of Sports Medicine (ACSM) ^6,24^, as well as national recommendations from the Dutch Health Council ^25^ and the Guideline Oncological Rehabilitation ^26^, recommend both cancer patients and healthy individuals to engage in at least 150 minutes of moderate-intensity aerobic activity per week, complemented by muscle-strengthening activities on two or more days per week. For cancer patients, these recommendations are supplemented with cancer-specific information that emphasizes the need to tailor physical activity to individuals’ abilities. Although international and national guidelines provide general recommendations for physical activity in cancer patients, it is unclear whether these are sufficiently applicable to glioma patients or if additional guidance is needed ^15^.

Given the limited glioma-specific evidence and the need to adapt general guidelines to individuals’ abilities, it is important to gain insight into how physical activity is currently integrated into the care of glioma patients. This understanding is not only important for informing clinical practice but also for guiding the development and evaluation of targeted physical activity interventions for this patient group. In this context, the perspective and role of healthcare professionals are particularly important. Healthcare professionals play a key role in promoting physical activity among cancer patients, including those with brain tumors, as their advice and support are important determinants of patients’ physical activity behavior ^27,28^.

Previous studies have explored the perspectives of oncology healthcare providers on promoting exercise in patients with various types of cancer. For instance, physical activity is not always prescribed due to limitations in time, knowledge, and resources ^28,29^. Another study revealed that healthcare professionals had concerns regarding physical strain and exhaustion in cancer patients, as well as issues related to professionals’ workload, coordination, and the availability of information and exercise programs. Additionally, patients’ physical fitness level and interest in physical activity were found to be key factors in whether physical activity was discussed ^30^.

We aimed to explore the experiences, perspectives, and barriers faced by healthcare professionals in promoting physical activity among glioma patients. While the findings aim to contribute to a broader understanding of these issues, this study specifically focused on the situation in the Netherlands as a starting point. By identifying these factors, we aimed to provide insights that can guide the development of practical implementation strategies to better integrate physical activity into glioma care.

## Methods

### Study design and participants

Healthcare professionals involved in the care of glioma patients in the Netherlands, including those working in the field of neurology, neurosurgery, oncology, radiotherapy, nursing (including nurse practitioners), and (neuro)psychology, were asked to complete a one-time survey. Only healthcare professionals from outside Amsterdam UMC were included in this study to avoid peer pressure, as the research was conducted from this center. Ethics approval was obtained from the Amsterdam UMC Medical Ethics Review Committee (2024.1146) and the participants provided online informed consent before participating.

### Survey

The anonymous survey comprised 19 questions in Dutch and took participants about 10-15 minutes to complete. The complete translated version of the survey is provided in supplementary table 1, following the original order in which the questions were presented. The survey was developed based on own experiences in the field and expert input, and was pilot-tested among a small group of researchers and professionals to ensure clarity. The survey included questions about the role of physical activity in the care of glioma patients (e.g. whether physical activity is discussed and what barriers exist), the potential effects of physical activity for glioma patients (e.g. advantages and disadvantages of physical activity), and physical activity guidelines (e.g. familiarity with physical activity guidelines and the need for specific recommendations). In addition to these questions, we collected characteristics of the participating professionals, including age, gender, specialty, years of experience, workplace, and professionals’ own physical activity behavior.

The survey consisted of 12 closed-ended questions and 7 open-ended questions. Closed-ended questions were scored on 5-point Likert scales (e.g. from ‘very often’ to ‘never’, ‘no barriers at all’ to ‘many barriers’, and ‘totally agree’ to ‘completely disagree’) or had a binary ‘yes’ or ‘no’ answer.

### Data collection

We developed the online survey using Castor’s electronic data capture ^31^ and distributed it via an open survey link. In February 2025, the survey was distributed via email by the Dutch Neuro-Oncology Society [Landelijke Werkgroep Neuro-Oncologie (LWNO)], the official association of physicians, nurses, and researchers working in the field of neuro-oncology and involved in the care or study of patients with tumors of the nervous system in the Netherlands ^32^. Additionally, we asked individual healthcare professionals from different centers to share the survey with their colleagues and asked the LWNO to send a reminder to all recipients in March 2025.

### Statistical analysis

The analyses were performed using R, version 4.2.3. Descriptive statistics, including frequencies and percentages, were used to analyze the closed-ended survey responses. Open-ended responses were systematically reviewed to identify recurring themes. Text fragments directly addressing the survey questions or providing meaningful insights were considered relevant and coded accordingly. Responses with similar content or addressing the same underlying idea were grouped into overarching thematic categories to capture recurring patterns and shared experiences across participants.

To enhance the reliability of the thematic analysis, two researchers (MB & MG) independently reviewed and coded the open-ended responses. Any discrepancies in coding or interpretation were resolved through discussion. In case of missing data, such as unanswered questions, the available responses were still included in the analysis.

## Results

### Characteristics of healthcare professionals

Table 1 summarizes the characteristics of the participants. A total of 55 healthcare professionals from 20 different centers completed the survey, with neurology and nursing being the best represented specialties. Most participants were women (74.5%), and years of experience in the field ranged from 0.5 to 29 years. The majority of participants reported engaging in regular and varied physical activity themselves, with frequencies ranging from several times a week to daily. One participant completed the closed-ended questions but skipped the open-ended questions. Apart from this, there was no missing data.

**Table 1.** Healthcare professionals characteristics.

| Characteristic | N = 55 |
| --- | --- |
| Age, mean (SD), years | 45.3 (9.90) |
| Gender, n (%) |  |
| Men | 14 (25.5) |
| Women | 41 (74.5) |
| Specialty, n (%) |  |
| Neurology | 19 (34.5) |
| Nursing | 18 (32.7) |
| Of whom nurse practitioners | 13 (24.1) |
| Radiotherapy | 8 (14.8) |
| Neurosurgery | 5 (9.1) |
| Oncology | 4 (7.3) |
| Psychology | 1 (1.8) |
| How many years working in this specialty, median (range) | 10 (0.5-29) |
| Centers included, n | 20 |

### Patient questions and professionals recommendations

The majority of healthcare professionals reported receiving questions about physical activity either (very) often (58.2%) or sometimes (36.4%), with none reporting never receiving such questions. The most frequently asked questions related to permission, practical advice, and the advantages of physical activity. These questions arose at all stages of the disease, with most occurring during treatment. An overview of survey questions and responses is provided in table 2.

**Table 2.**
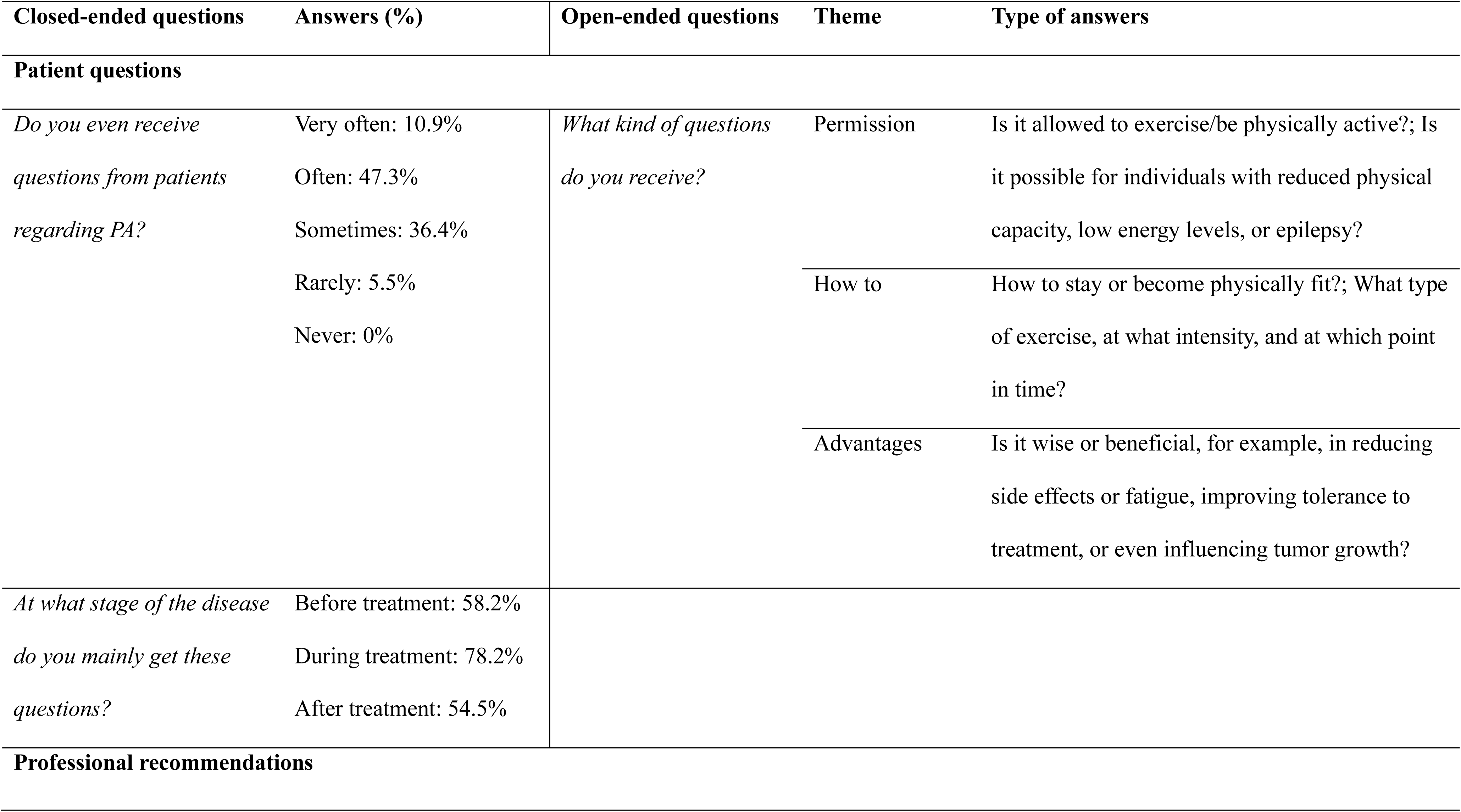
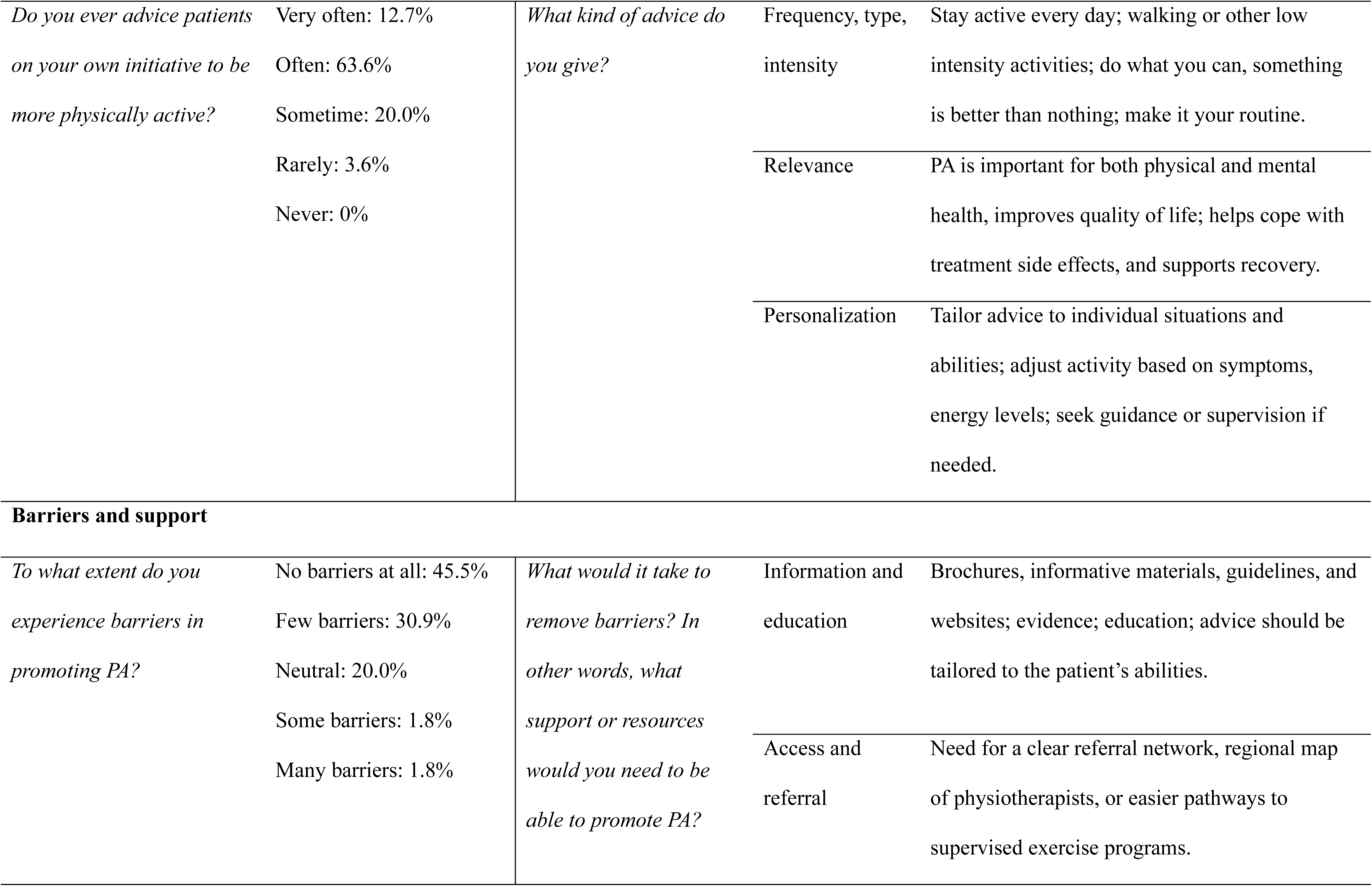
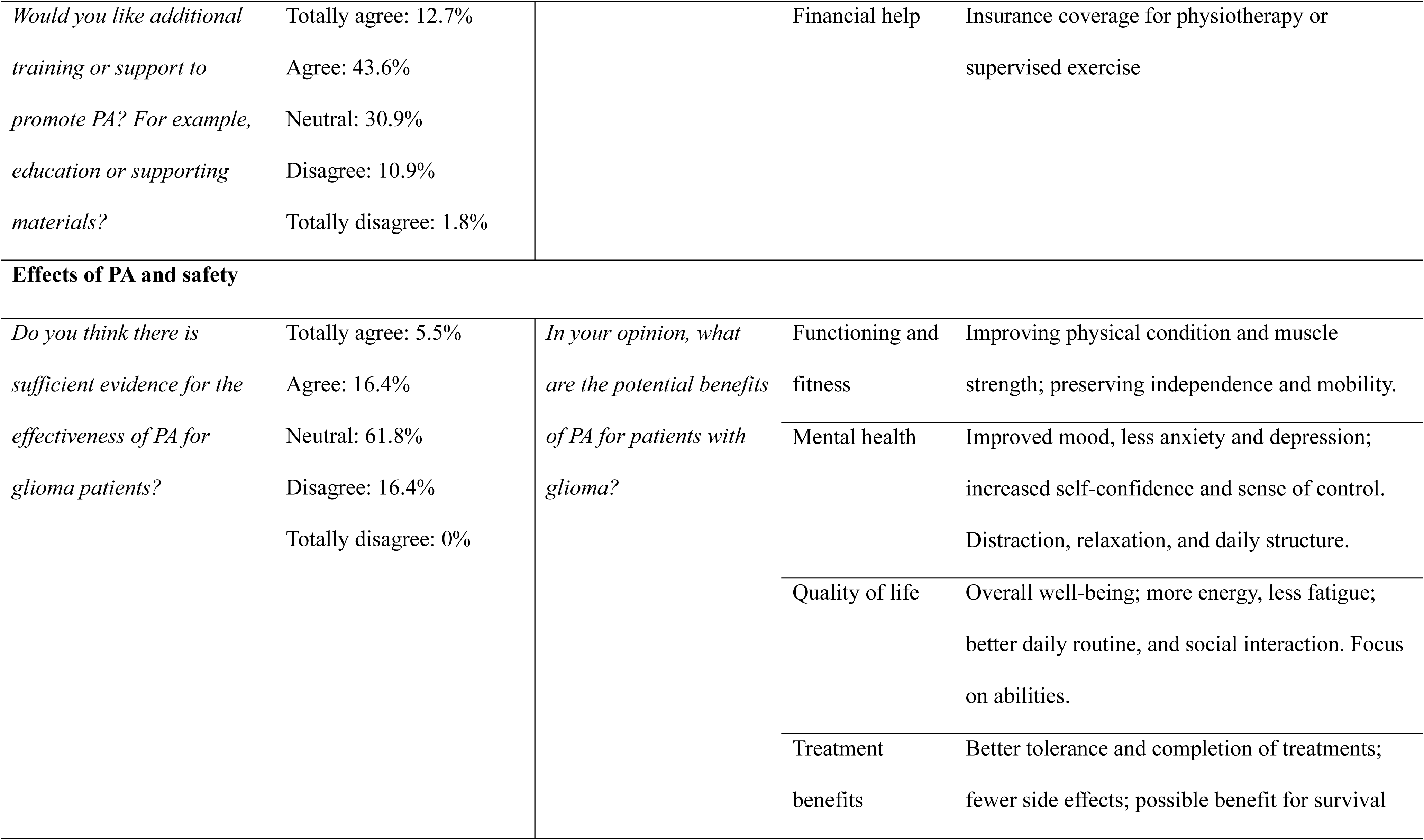
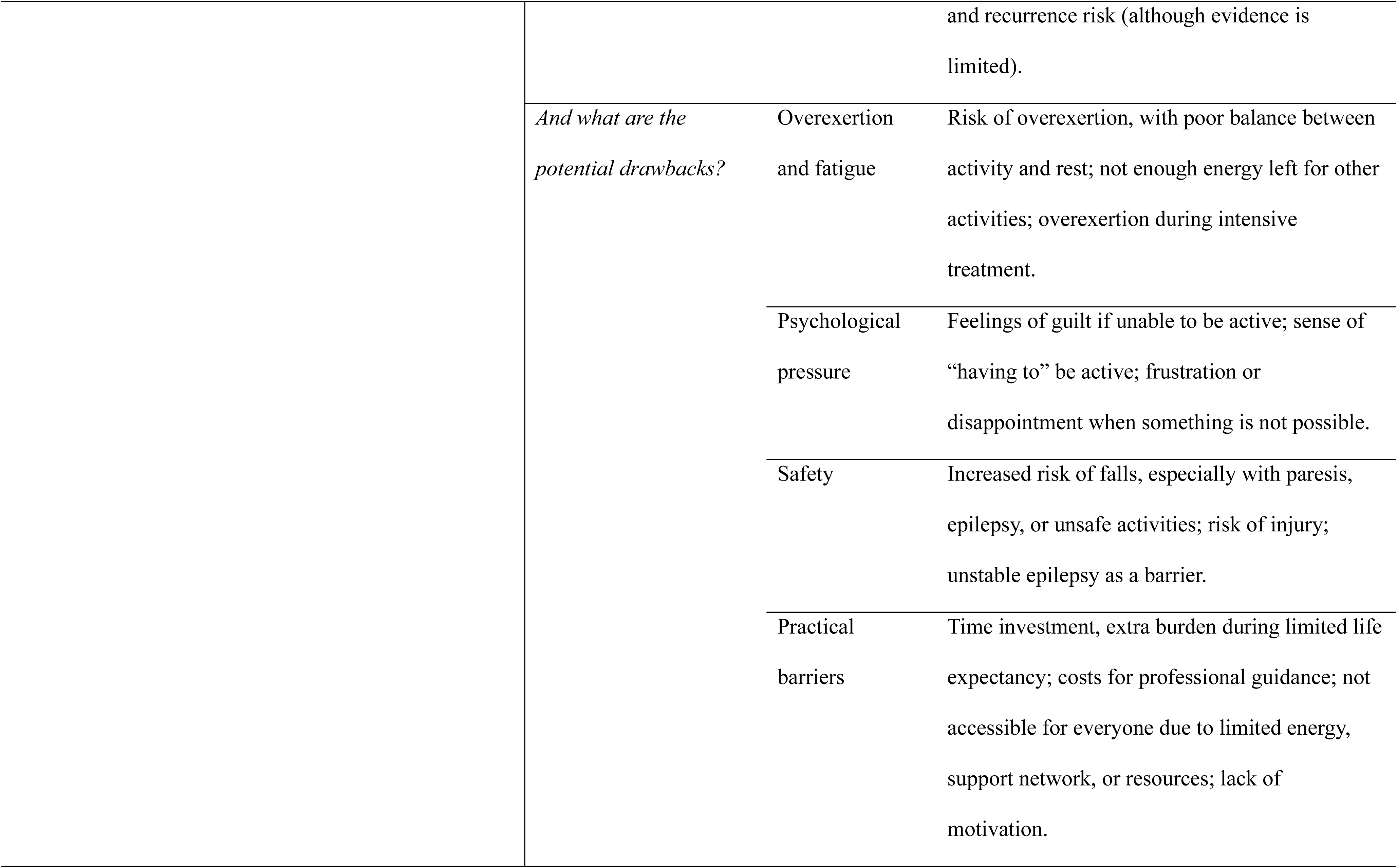
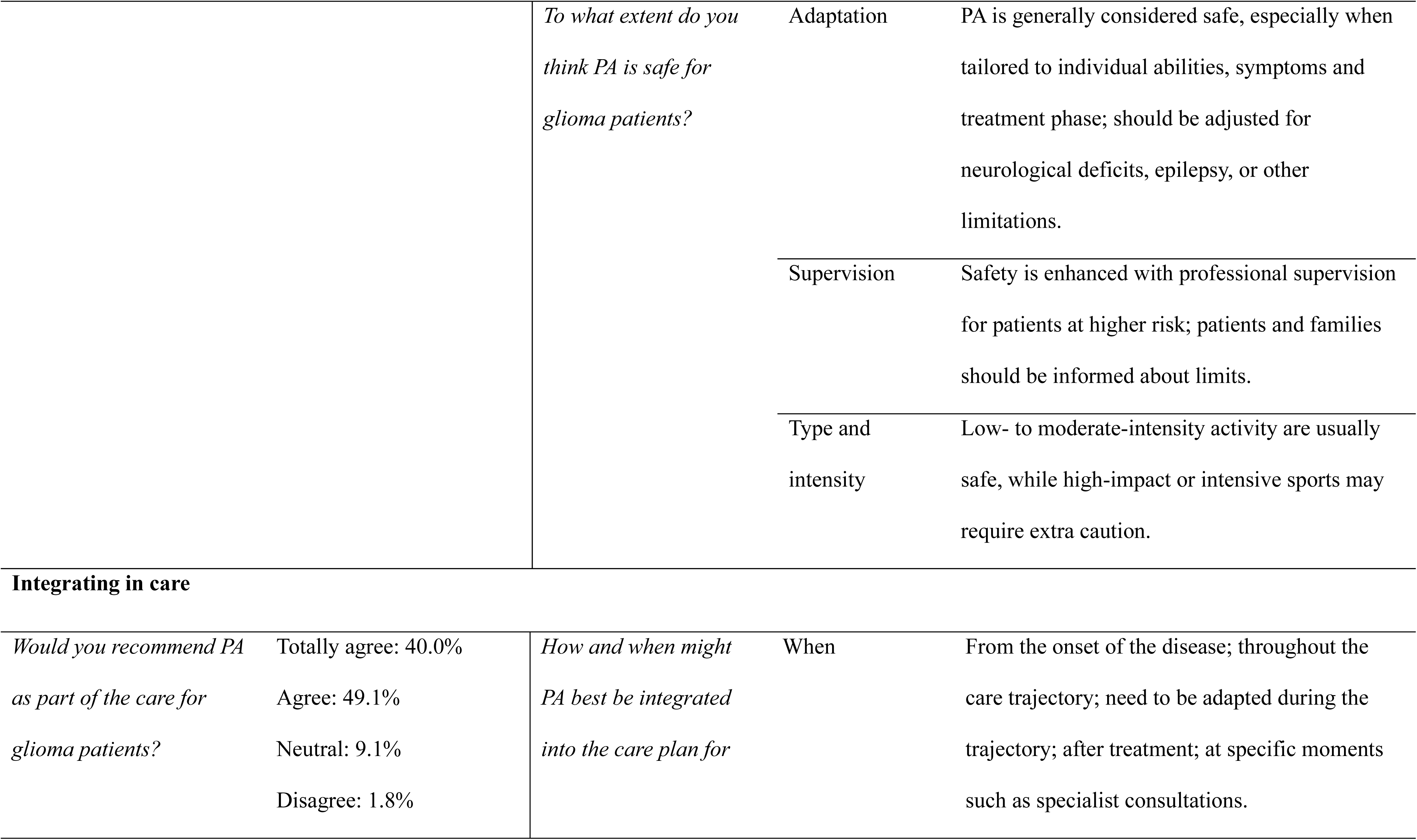
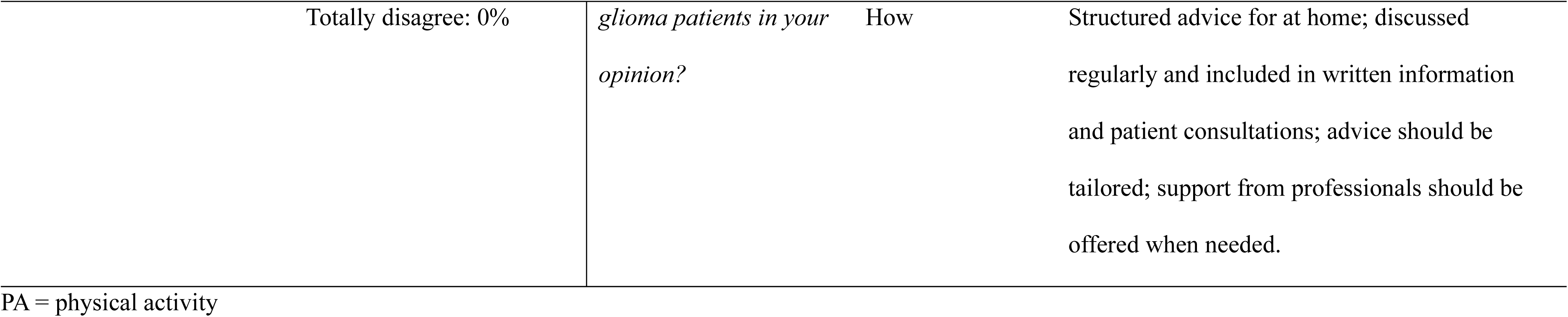
Thematic overview of survey questions and answers.

The majority of healthcare professionals reported that they (very) often (76.3%) proactively offered recommendations on physical activity. These mainly focused on encouraging daily activity, highlighting the importance of PA for both physical and mental health, and tailoring advice to the individual needs and abilities of each patient.

### Experienced barriers and need for support

Nearly half of the healthcare professionals (45.5%) indicated not experiencing any barriers in recommending or promoting physical activity, while the others reported experiencing barriers to varying degrees. The healthcare professionals who reported experiencing barriers (54.5%) were asked to specify these barriers (*Fig. 1*). Commonly mentioned barriers included limited informational material, limited knowledge, limited knowledge of, and uncertainty about available referral options, and time constraints. Barriers reported by healthcare professionals under the ‘Other’ option included patients’ financial difficulties due to lack of insurance coverage, the challenge of recommending physical activity to patients who are severely affected by their disease, limited knowledge about available services in the region, and the need to avoid overwhelming patients with too much information.

**Figure 1.**
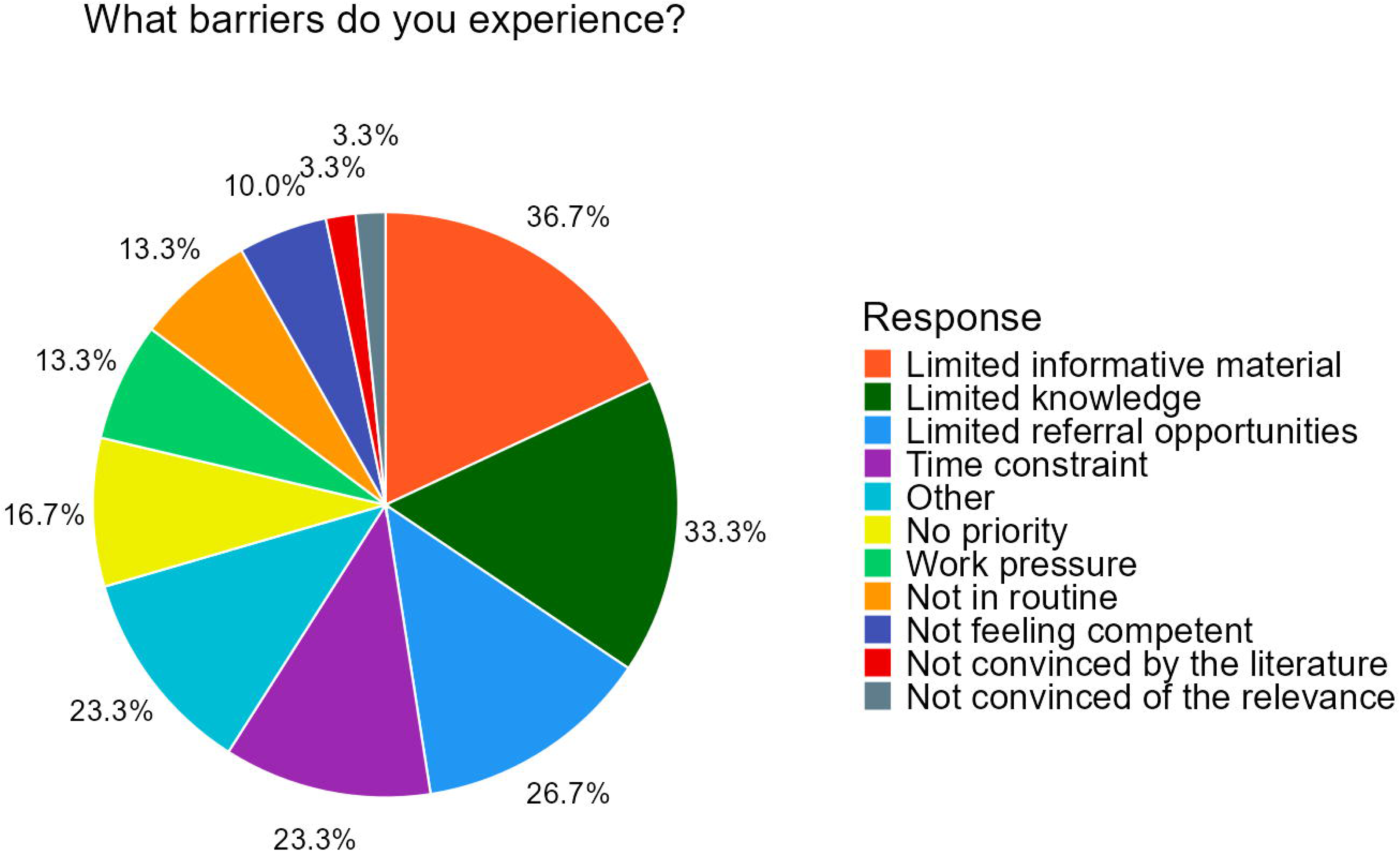
Responses from healthcare professionals regarding the types of barriers encountered.

To remove barriers and better promote physical activity, healthcare providers often mentioned the need for informational materials or brochures, as well as established referral pathways or access to specialized physiotherapists. An overview of survey questions and responses is provided in table 2. The majority (56.3%) reported they would need additional training or support to be able to promote physical activity, 30.9% were neutral, while 12.7% (totally) disagreed.

### Perceived effects of physical activity and safety

Commonly reported potential benefits of physical activity by healthcare professionals were improved fitness and functioning, a positive effect on mental well-being, improved quality of life, better ability to cope with treatment, and fewer side effects. An overview of survey questions and responses is provided in table 2. Commonly reported potential drawbacks included the risk of fatigue or overexertion, psychological pressure, safety concerns, and practical barriers. Almost all healthcare professionals (totally) agreed that physical activity can help reduce symptoms (96.4%; *Fig. 2A*) and improve quality of life (98.2%; *Fig 2B*). Healthcare professionals also agreed that physical activity is generally safe for glioma patients, as long as it is appropriately adapted to the individual’s condition.

**Figure 2.**
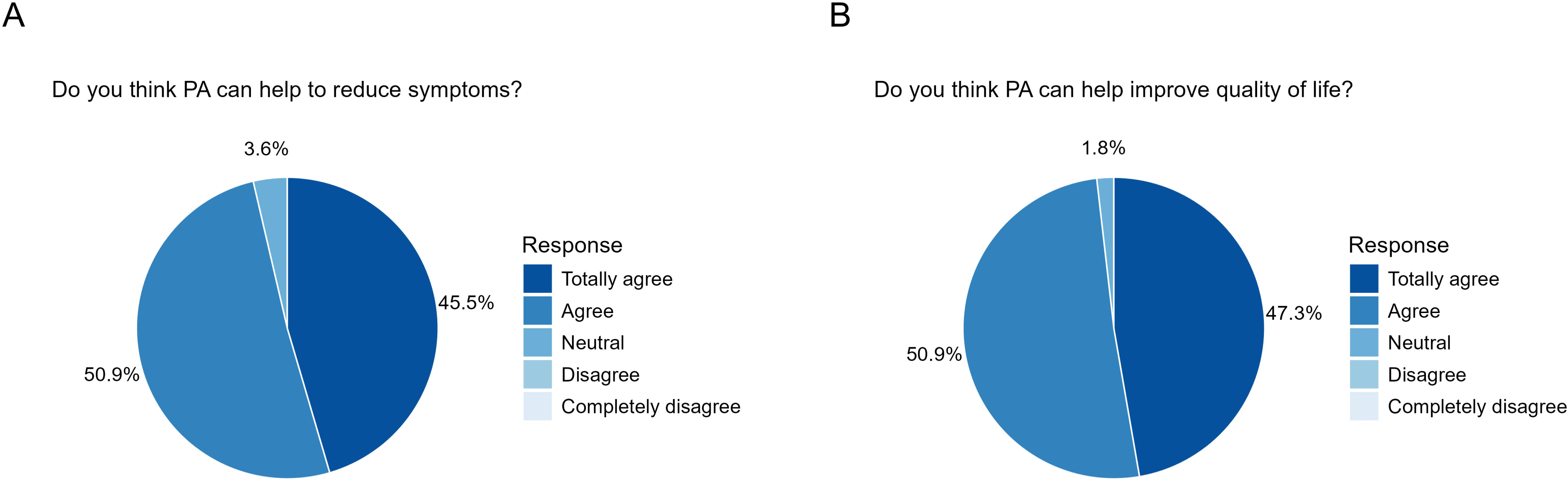
Healthcare professionals’ perceptions of the potential of physical activity (PA) to reduce symptoms such as fatigue (**A**) and improve quality of life (**B**).

Perspectives varied more regarding whether there is sufficient scientific evidence for the effectiveness of physical activity, with 61.8% of healthcare professionals indicating a neutral view and 16.4% disagreeing.

### Integration of physical activity into care

The majority of healthcare professionals (89.1%) agreed or completely agreed that physical activity should be part of the care for glioma patients. An additional 9.1% were neutral, and only one healthcare professional disagreed.

In response to the question of when and how physical activity should be integrated into the care plan of glioma patients, the majority of healthcare professionals indicated that this should begin at diagnosis or at the start of treatment and be maintained throughout the entire care trajectory. They also emphasized the importance of a patient-centered approach. An overview of survey questions and responses is provided in table 2.

### Familiarity with physical activity guidelines

The majority of healthcare professionals indicated they were familiar with the general physical activity guidelines (*Fig. 3A*) and reported that they feel the need for a guideline specific to glioma patients (*Fig. 3B*).

**Figure 3.**
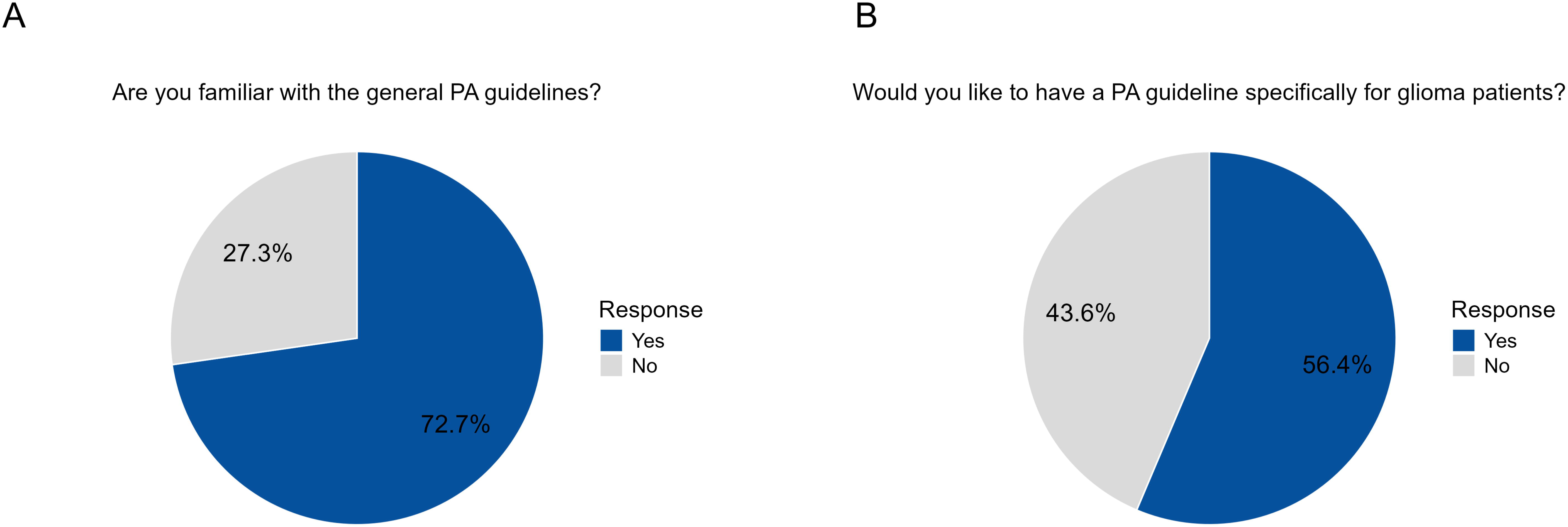
Healthcare professionals’ (**A**) familiarity with general physical activity (PA) guidelines and (**B**) perceived need for PA guidelines specifically tailored to patients with glioma.

### Perceived barriers and support needs across professional groups

To gain further insight, we performed post hoc analyses exploring barriers and support needs across different professional groups, levels of experience, and attitudes towards guidelines.

Among the healthcare professionals who reported rarely or sometimes providing proactive physical activity recommendations (N = 13), most experienced few (46.2%) or some barriers (38.5%), while only a small proportion reported no barriers (15.4%, *Supplementary* Fig. 1A). However, this group of professionals did not fully align with those who expressed a need for additional support, as a relatively larger percentage (23.1%) disagreed with requiring further training or support (*Supplementary* Fig. 1B). We found no difference between neurologists and nurses compared to other specialties in how often they proactively recommend physical activity.

The need for additional training or support to be able to promote physical activity was more frequently expressed by professionals with over 10 years of experience in the field, with 69.2% agreeing or totally agreeing (*Supplementary* Fig. 2A), compared to 44.8% among those with 10 years or less experience (*Supplementary* Fig. 2B). In contrast, the difference in reported barriers between the two groups with different seniority in the field was less pronounced, although healthcare professionals with 10 years or more of experience reported slightly more barriers (*Supplementary* Fig. 2C*, 2D*).

Among healthcare professionals who indicated no need for specific guidelines for glioma patients, the majority (62.5%) experienced no barriers at all to promoting physical activity (*Supplementary* Fig. 3A). In addition, a smaller percentage reported needing support to promote physical activity, compared to all healthcare professionals combined (*Supplementary* Fig. 3B).

## Discussion

We aimed to explore the experiences, perspectives, and barriers faced by healthcare professionals in promoting physical activity among glioma patients. Our findings show that healthcare professionals frequently receive questions about physical activity, acknowledge its importance, and proactively give recommendations on this subject. Nevertheless, notable barriers persist, such as limited informative materials, lack of knowledge, and limited referral options. Additionally, the quality of the advice given could be improved. These perceived limitations highlight broader challenges in the promotion of physical activity for glioma patients.

While the benefits of physical activity are well established in the general oncology population, evidence specifically supporting the effectiveness of physical activity interventions in glioma patients remains limited. Addressing this lack of targeted research will contribute to the ability to provide evidence-based, tailored advice for this patient group. At the same time, even the general physical activity guidelines that are available are not integrated into clinical practice, and the recommendations currently provided to glioma patients are often limited in scope and specificity. Addressing both the evidence gap and the practical barriers is essential for improving the support and guidance offered to glioma patients regarding physical activity.

Prior research focusing on exercise interventions for brain tumor patients has shown promising results, particularly demonstrating safety and feasibility, but also indicating positive effects on aerobic capacity, physical functioning, psychological well-being, and reductions in fatigue and depression ^9,10,13,33-36^. While nearly all healthcare professionals in this study believe that physical activity has the potential to improve symptoms like fatigue and enhance quality of life, and would recommend it as part of care, the majority remain uncertain whether there is enough evidence to fully support this recommendation.

These challenges and uncertainties directly influence the way healthcare professionals give recommendations to glioma patients about physical activity. Healthcare professionals in this study encourage glioma patients to be physically active every day, focusing on low-intensity activities such as walking, and emphasize that any activity is better than no activity. However, current general and cancer-specific physical activity guidelines promote a more structured and intensive approach ^6,23-25^. Cancer-specific guidelines ensure that physical activity remains safe and effective throughout the disease trajectory by addressing treatment-related side effects and fluctuations in physical capacity ^37^. It remains uncertain whether these general guidelines are fully appropriate for this patient group ^37-39^. Compared to the general cancer population, brain tumor patients face distinct challenges such as neurological and cognitive impairments, and an increased risk of epileptic seizures ^3^. As specific guidelines for glioma patients are currently limited ^15,40^, it may be most appropriate for healthcare professionals to use existing cancer-specific guidelines as a flexible framework, adapting them to the individual challenges of glioma patients. As more tailored evidence becomes available, these recommendations can be further refined.

In addition to addressing professional perspectives, it is also important to consider the preferences and needs of patients themselves. A previous study found that patients with a primary brain tumor have varying needs and preferences for exercise programs, which should be tailored to their treatment phase and physical condition ^41^. Such a study could provide valuable insights into patients’ preferences, helping to ensure that future physical activity programs are well aligned with the needs of glioma patients. Future research should continue to explore these preferences.

The questions most frequently received by healthcare professionals were broad, including whether exercise is permitted, which activities are appropriate, and what the potential benefits are. These questions often focused on the existence of specific recommendations. This suggests that, in addition to healthcare professionals, patients also have a need for information presented in the form of clear guidelines. In addition, these questions were asked throughout all stages of the disease, with a slight increase during the treatment phase. This may be explained by previous research indicating that physical activity levels are lowest during treatment ^15^, leading to a greater need for information at this time.

The barriers identified in this study largely correspond with those found in other cancer populations, such as lack of time, limited access to trained specialists or referral options, not feeling qualified to discuss or refer to exercise, concerns about overexertion and psychological stress for patients ^28-30^. Additionally, high workloads and limited information further complicate the promotion of physical activity ^29^. As a result, healthcare professionals tend to promote physical activity selectively and cautiously, mainly to patients who are already active rather than those who are sedentary ^30^. These findings indicate that the challenges faced in promoting physical activity among glioma patients are not unique, but reflect broader issues observed across oncology care.

One limitation of this study is uncertainty regarding the representativeness of the sample, which may limit the generalizability of our findings. In addition, selection bias may have occurred due to the use of an open survey link, potentially attracting healthcare professionals with a particular interest in physical activity. To explore this, we assessed the participants’ physical activity behavior, which indicated that the group was generally quite active. Additionally, self-reported responses may have been influenced by social desirability bias. This may have led participants to provide more favorable answers than they would in clinical practice.

In summary, nearly all healthcare professionals in this study acknowledged the value of physical activity in reducing symptoms and improving quality of life in glioma patients, indicating a strong consensus within the field. While physical activity is frequently discussed in glioma care, the recommendations provided could be improved. Developing specific physical activity guidelines for glioma patients could address common barriers and support more effective promotion of physical activity in clinical care. Future research should further evaluate the effects of physical activity interventions for glioma patients to inform and optimize clinical practice.

## Ethics

Approval for the collection of the data was obtained from the Amsterdam UMC Medical Ethics Review Committee (2024.1146).

## Authorship

Conceptualization and design: MB, LD, MK, MG, PdwH, AN, JJ

Data collection and curation: MB, LD, MK

Data analysis and interpretation: MB, LD, MK, MG

Visualization: MB, LD, MK

Writing the manuscript: MB, MK, LD

Revising of the manuscript: All authors

Final approval of manuscript: All authors

Accountable for all aspects of work: All authors

## Data availability

Data will be made available upon reasonable request.

