## Supplementary materials for "Promoting physical activity in glioma patients: insights from Dutch healthcare professionals"

**Supplementary table 1. English translation of the original Dutch survey**

| **Healthcare professional profile** | | |
|---|---|---|
| *Question* | | *Answer (open or closed)* |
| 1 | What is your gender? | o Male<br>o Female<br>o Non-binary<br>o Other, namely |
| 2 | What is your age? | |
| 3 | What is your specialty? | o Neurologist<br>o Neurosurgeon<br>o Oncologist<br>o Radiotherapist<br>o Nurse<br>o Nurse specialist<br>o Rehabilitation physician<br>o Physiotherapist<br>o (Neuro)psychologist<br>o Other, namely: |
| 4 | How many years have you been working in this specialty? | |
| 5 | In which center do you work? | |

| **Current care of glioma patients** | | |
|---|---|---|
| *Question* | | *Answer (open or closed)* |
| 6 | Do you ever receive questions from patients regarding physical activity/exercise/sports/fitness? | Never – rarely – sometimes – often – very often |
| 7 | What kind of questions do you receive? | |
| 8 | At which stage of the disease do you mainly receive these questions? | *Multiple answers possible*<br>o Before treatment<br>o During treatment<br>o After treatment<br>o Other, namely: |
| 9 | Do you ever advise patients on your own initiative to be more physically active? | Never – rarely – sometimes – often – very often |
| 10 | If so, what kind of advice do you give? | |
| 11 | To what extent do you experience barriers in giving physical activity advice/promoting physical activity? | No barriers at all – few barriers – neutral – some barriers – many barriers |
| 12 | What barriers do you experience? | *Multiple answers possible*<br>o Not feeling competent<br>o Limited knowledge<br>o Workload<br>o Not part of routine<br>o Time<br>o Availability of informative material |

| | | |
|---|---|---|
| | | ○ Not convinced by the literature<br>○ Not convinced of the relevance<br>○ Not a priority<br>○ Limited referral options<br>○ Other, namely: |
| 13 | What is needed to remove these barriers? In other words, what support or resources would you need to be able to give physical activity advice? | |
| 14 | Would you like additional training or support to promote physical activity in glioma patients? For example, education or supporting materials. | Strongly agree – agree – neutral – disagree – strongly disagree |
| 15 | In your opinion, how and when should physical activity best be integrated into the care plan for glioma patients? | |

| **Physical activity in glioma patients** | | |
|---|---|---|
| *Question* | | *Answer (open or closed)* |
| 16 | What do you think are the potential benefits of physical activity for patients with glioma? | |
| 17 | And what are the potential disadvantages? | |
| 18 | Do you think physical activity can help reduce symptoms (such as fatigue, pain, anxiety, etc.)? | Strongly agree – agree – neutral – disagree – strongly disagree |
| 19 | Do you think physical activity can help improve quality of life? | Strongly agree – agree – neutral – disagree – strongly disagree |
| 20 | To what extent do you consider physical activity safe for glioma patients? Can you elaborate on your answer? | |
| 21 | Do you think there is sufficient evidence for the effectiveness of physical activity for glioma patients? | Strongly agree – agree – neutral – disagree – strongly disagree |
| 22 | Would you recommend physical activity as part of the care for glioma patients? | Strongly agree – agree – neutral – disagree – strongly disagree |

| **Physical activity guidelines** | | |
|---|---|---|
| *Question* | | *Answer (open or closed)* |
| 23 | Are you familiar with the general physical activity guidelines (for the general population)? | ○ Yes<br>○ No |
| 24 | Would you like to have a physical activity guideline specifically for glioma patients? | ○ Yes<br>○ No |
| 25 | What do you do yourself in terms of physical activity in a week? In what form and how often? | |

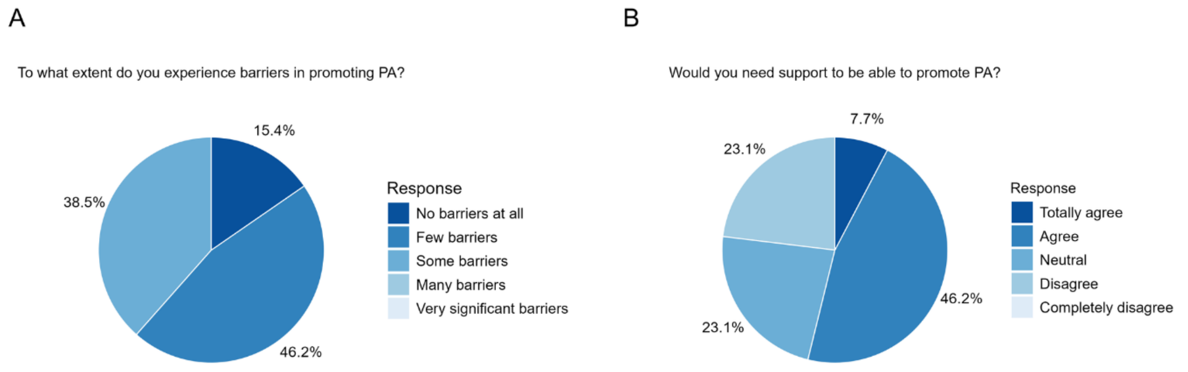


**Supplementary figure 1.** (**A**) Perceived barriers among healthcare professionals who rarely or sometimes provide proactive physical activity advice (N=13). (**B**) Perceived need for additional training or support in the same group.

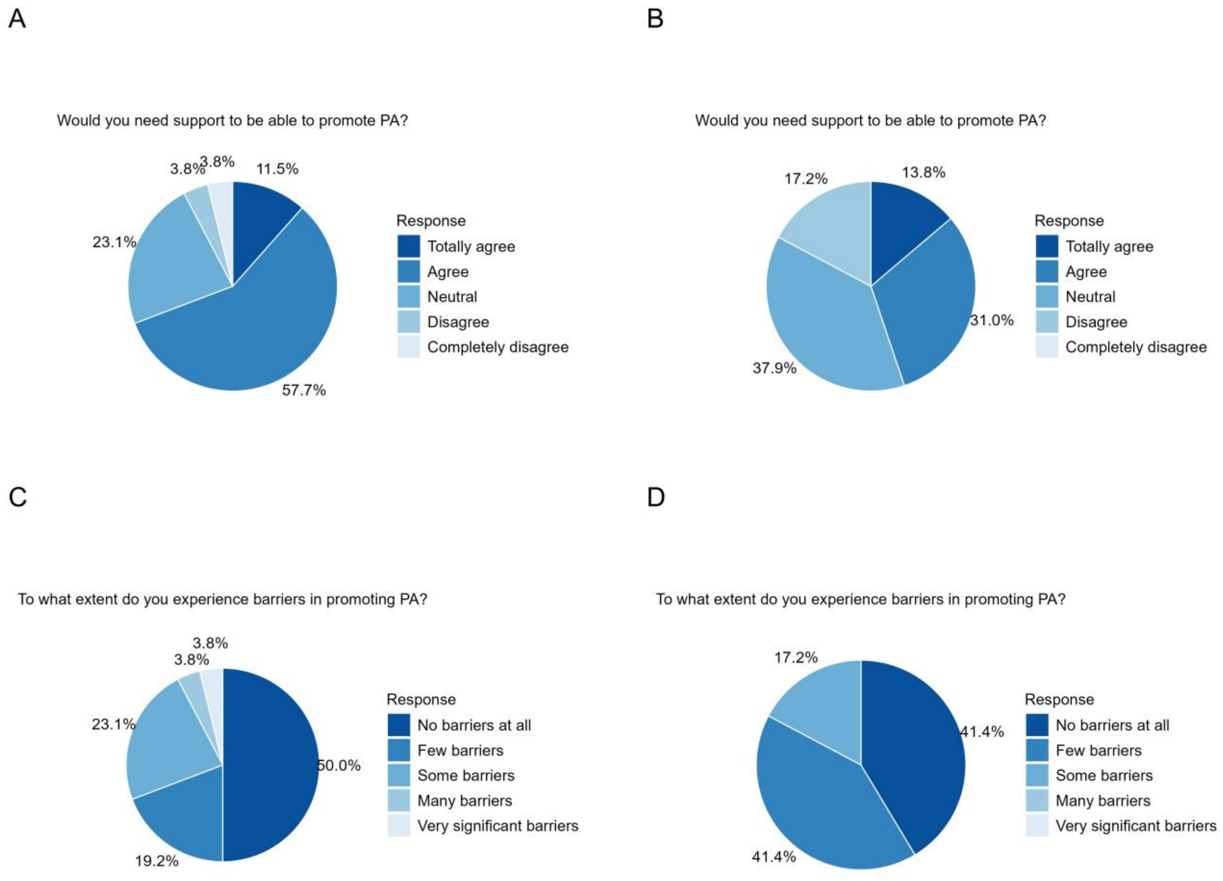


**Supplementary figure 2.** Perceived need for additional training or support to promote physical activity among healthcare professionals with more than 10 years of experience (N = 26; **A**) and 10 years or less (N = 29; **B**). Perceived barriers to promoting physical activity among healthcare professionals with more than 10 years of experience (N = 26; **C**) and 10 years or less (N = 29; **D**).

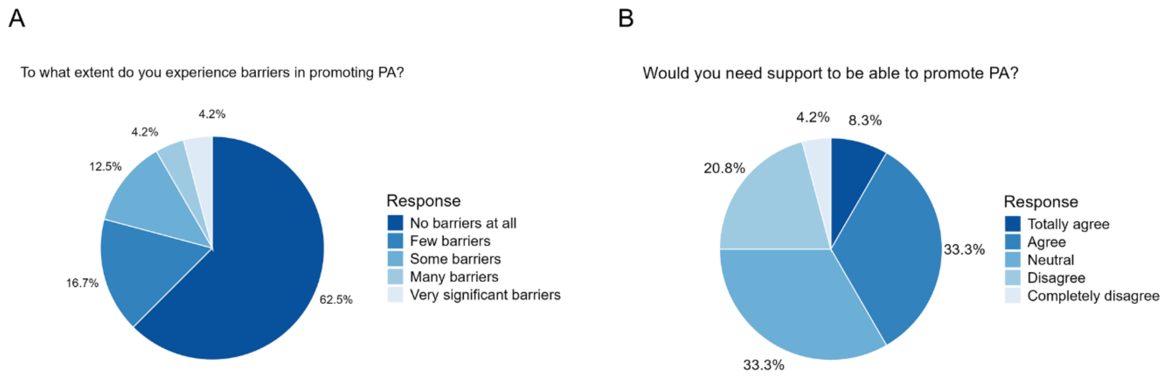


**Supplementary figure 3.** (**A**) Perceived barriers among healthcare professionals who indicated no need for specific guidelines for glioma patients. (**B**) Perceived need for additional training or support in the same group.
